# Evaluation of COVID-19 RT-qPCR test in multi-sample pools

**DOI:** 10.1101/2020.03.26.20039438

**Authors:** Idan Yelin, Noga Aharony, Einat Shaer Tamar, Amir Argoetti, Esther Messer, Dina Berenbaum, Einat Shafran, Areen Kuzli, Nagam Gandali, Tamar Hashimshony, Yael Mandel-Gutfreund, Michael Halberthal, Yuval Geffen, Moran Szwarcwort-Cohen, Roy Kishony

## Abstract

The recent emergence of SARS-CoV-2 lead to a current pandemic of unprecedented levels. Though diagnostic tests are fundamental to the ability to detect and respond, many health systems are already experiencing shortages of reagents associated with this test. Here, testing a pooling approach for the standard RT-qPCR test, we find that a single positive sample can be detected even in pools of up to 32 samples, with an estimated false negative rate of 10%. Detection of positive samples diluted in even up to 64 samples may also be attainable, though may require additional amplification cycles. As it uses the standard protocols, reagents and equipment, this pooling method can be applied immediately in current clinical testing laboratories. We hope that such implementation of a pool test for COVID-19 would allow expanding current screening capacities thereby enabling the expansion of detection in the community, as well as in close integral groups, such as hospital departments, army units, or factory shifts.

## Introduction

The ongoing pandemic of the recently-emerged SARS-CoV-2 is critically challenging health systems worldwide. The virus is characterized by fever and severe acute respiratory syndrome [1,2]. As of March 17, the World Health Organization (WHO) has reported over 170,000 cases with over 10,000 new diagnoses added in 24 hours [3].

Detecting carriers of the virus is fundamental to response efforts. It ensures the quarantine of COVID-19 patients to prevent local spread [1], and more broadly informs national response measures [4]. Nevertheless, as monitoring capacity is limited, testing in most countries is generally focused on acutely ill patients, while potentially infectious carriers at the community remain undiagnosed. As many countries are already experiencing shortages of diagnosis kits and factories struggling to keep with the demand [5–7], it has become important to come up with new ways to conserve the reagents used for diagnostic tests. At the same time, as the disease is novel, it is of value to validate any modifications to the testing process before universal adoption [8,9].

Pooling diagnostic tests has been applied in other infectious diseases and is especially attractive as it requires no additional training, equipment, or materials. In this method, first suggested by Dorfman in 1943 [10] and perfected over the years [11–13], samples are mixed and tested at a single pool, and subsequent individual tests are made only if the pool tests positive. In addition to being used in the clinic for infectious disease diagnostics in previous epidemics [14,15], pooling has been proven to work for RT-qPCR [16,17], a time-consuming step for which the reagents are expected to be in short supply [18]. Nonetheless, as SARS-CoV-2 is a novel pathogen, it is unclear how diluting a sample containing its RNA would affect the sensitivity of this assay and the false-negative rate.

Here, we test the ability of the standard RT-qPCR test for detecting a single positive sample within a pool of negative samples. Pooling clinical RNA samples, we tested previously confirmed positive samples alone and combined with an increasing number of previously confirmed negative samples and found that positive samples can still be well observed in pools of up to 32 samples, and possibly even 64 with additional PCR cycles.

## Methods

### Sample collection

Swabs from both nostrils and the throat were previously collected by healthcare providers and sent to the Virology laboratory at the Rambam Health Care Campus, Haifa, Israel. A volume of 130 microliter of the transport swab buffer was mixed with 270 microliter lysis buffer and RNA was extracted using magLEAD (Precision System Science). We obtained samples tested between March 4-15, 2020.

### Individual RT-qPCR tests in the clinical laboratory

RT-qPCR was performed in the clinical laboratory to detect the presence of SARS-CoV-2 RNA with AgPath-ID™ One-Step RT-PCR Reagents (Thermo Fisher Scientific) in a Bio-Rad CFX 96 qPCR machine with WHO primers and probe (E_Sarbeco_R: ATATTGCAGCAGTACGCACACA, E_Sarbeco_F: ACAGGTACGTTAATAGTTAATAGCGT, E_Sarbeco_P: ACACTAGCCATCCTTACTGCGCTTCG) [19]. Reactions were heated to 50°C for 30 minutes for reverse transcription, denatured in 95°C for 10 minutes and then 46 cycles of amplification were carried in 95°C for 15 seconds and 55°C for 32 seconds. Fluorescence was measured using the FAM parameters.

### Pooled-samples RT-qPCR in the research laboratory

Laboratory RT-qPCR procedure was performed according to the procedure for individual samples in the clinical laboratory, on an identical qPCR machine and program and with reagents donated from the Rambam Health Care Campus. To conserve resources and allow multiple pooling and duplicates of the same sample, each sample was diluted by X0.4 prior to mixing with reagents.

### Pooling

We arbitrarily chose 5 positive samples and 67 negative samples. 66 of the negative samples were mixed into 8 different pools containing equal volumes of 1, 2, 4, 8, 16, and 32 unique samples. Samples 1-3 varied between replicates to determine whether different negative-sample composition in the pool affected the detection of positive samples. The final 67th sample was mixed with the pool of negative samples as control for the positive samples. The negative pools were distributed in 6 rows of a 96-well plate, 5ul per well, and 10ul of the positive samples and the 67th negative sample were distributed in the 7th row. 5ul of the positive samples were then diluted into the “pool” of 1 sample to make a ½ dilution, then the ½ dilution was diluted in the 2 samples pool to make a 1/4 dilution etc., up to 1/64. Finally, 20ul of the RT-qPCR reagent mix were added to each well.

### Ethical approval

This study was granted exemption from IRB approval for use of deidentified discarded RNA samples of COVID-19 tests by The Rambam Health Care Campus IRB committee.

## Results

The original diagnostic run at the Rambam Health Care Campus was robust. Positive samples had on average 135 ± 32-fold stronger fluorescence relative to negatives, and the positive samples reached the threshold, which we set at a fluorescence of 300 according to CDC guidelines [8], at the 25.5± 6.1 cycle (Figure 1, Supplementary Figure 1). The five positive samples selected (marked above) similarly averaged at Ct of 24.5±-3.1 and maximum fluorescence of 5164±912.

**Figure 1:**
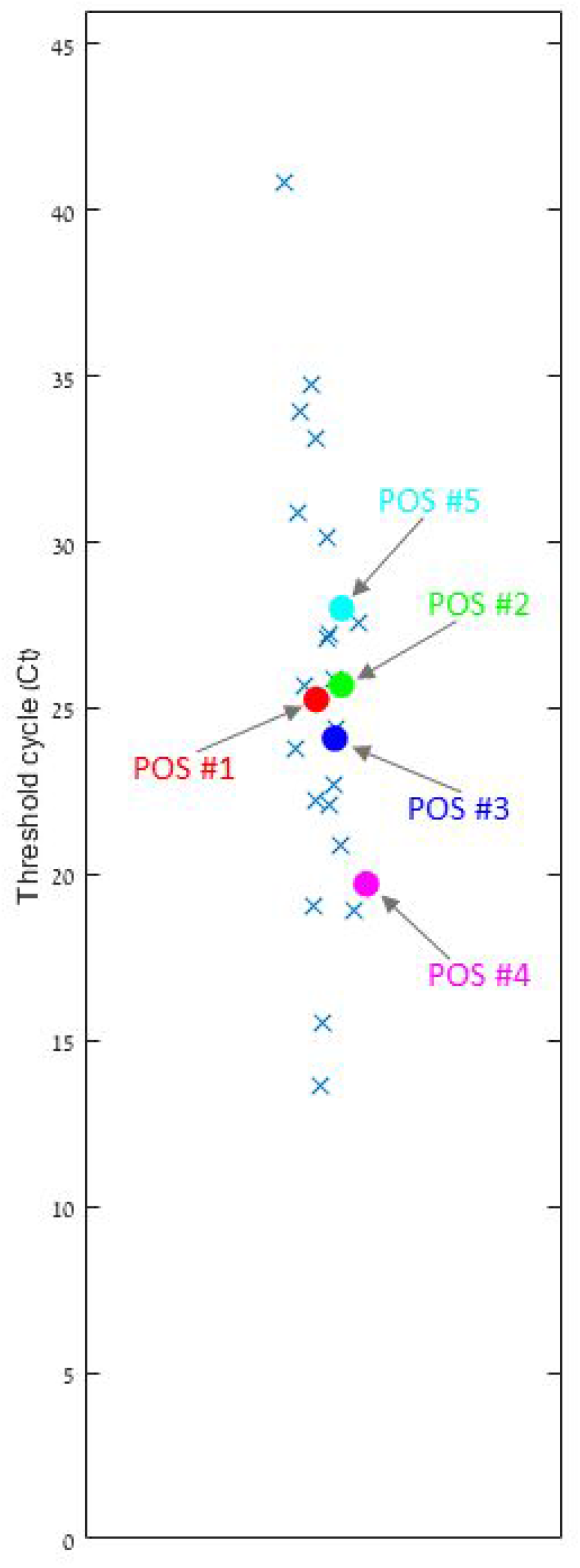
Selection of positive samples. Threshold cycle (Ct) of Positive Samples from Rambam Health Care Campus showing 26 positive samples (out of 388 samples tested) had an average Ct of 25.5+/- 6.1 (standard deviation). Out of these samples, we selected 5 representative positive samples marked POS #1-5 (colored circles).

**Figure 2:**
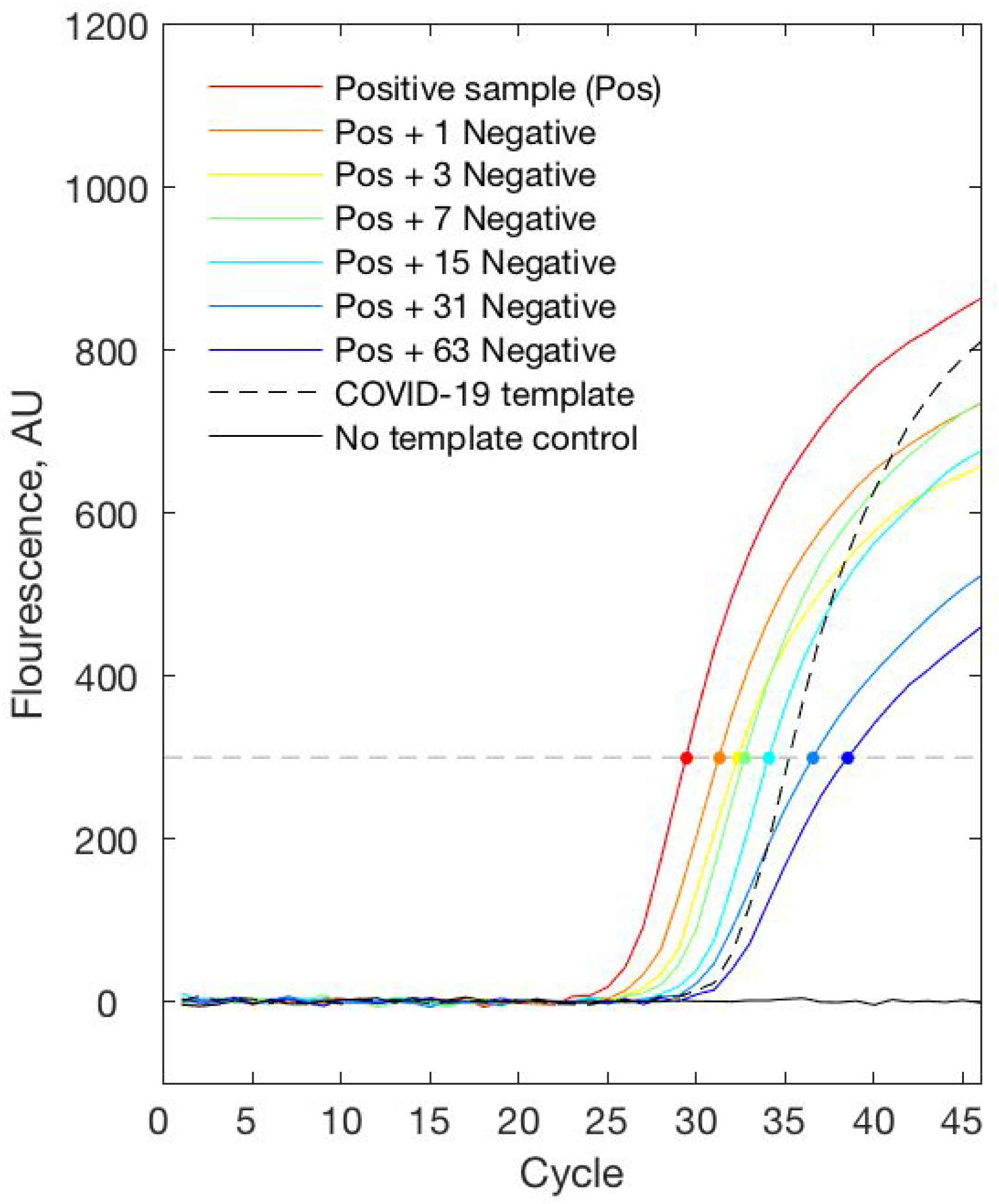
Replicate A of Positive sample #1 is detected when mixed with up to 63 negative samples. Representative RT-qPCR fluorescence curves of a positive sample (POS #1) diluted in different numbers of negative samples (red - no dilution, blue - dilution in 63 negative samples). Dots represent the cross point of the fluorescence threshold (threshold = 300, gray dashed line).

As the number of negative pooled samples increases, the amplified RNA reaches the threshold later, as expected from a diluted sample. Except for a single replicate (POS #2, supplementary Figure 2), all samples reached the threshold in 32-sample pools. For most samples there is a linear correlation between when the threshold is reached and the doubling of the pool size, corresponding with the expectation that an RNA sample that is diluted twice as much will take one more cycle to double in amount and reach the same fluorescence (Figure 3). The observed linearity indicates that in most cases there is no RNA interference with the reverse transcriptase or DNA polymerase enzyme.

**Figure 3:**
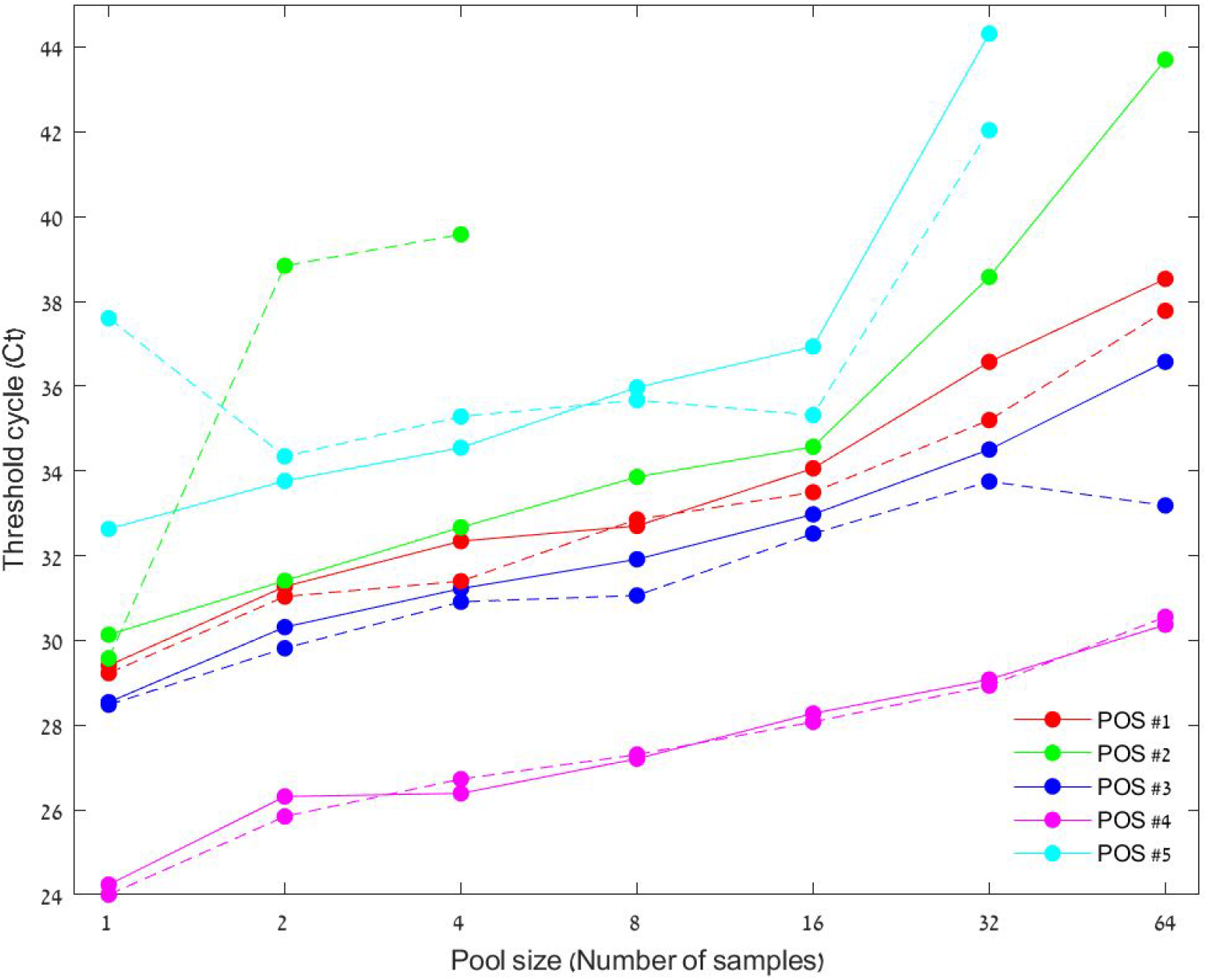
Positive samples are consistently detected when diluted with up to 31 negative samples. Pool size containing a single positive sample over the RT-qPCR cycle where it crosses the threshold (Filled line - duplicate A, dashed line - duplicate B). Most positive samples reach the threshold at a later Ct as they are more diluted. Samples #2 and #5, which reached the threshold later than others, grew nonlinearly relative to other samples.

Of the ten tested replicates, only replication B of sample #2 did not cross the threshold in pools of 32. Moreover, with the exception of this specific replicate, the fluorescence of all 64-sample pools increased in a sigmoidal manner. In contrast, negative samples in replication B in the 64-, 32-, and 2-sample pool also began to increase but none maintained a sigmoidal pattern or crossed the threshold (supplementary Figure 2).

## Discussion

We found that a single clinical sample with SARS-CoV-2 RNA can be consistently detected in a pool of up to 32 samples. Pooling this way leads to only a linear increase in the threshold cycle (Ct). Our data shows an estimated false negative rate of 10% (1 out of 10), which is relatively small compared to the inherent clinical sensitivity of the standard assay [20].

RT-qPCR could be further optimized for the detection of low-concentration RNA. For instance, additional amplification cycles could lower detection limit allowing better detection for pools of more than 32 samples, which based on extrapolation of the data we expect would allow the 64-sample pools of positive sample #5 to cross the threshold. In addition, some abnormalities as with duplicate B in positive samples #5 and #2 could have been due to interference from contamination in one or more of the samples. The unusual peaks for positive sample #2 could be due to a changing salt concentration that disturbed the TaqMan probe. Since both these issues could be solved by further diluting the RNA samples with water, it is worthwhile to explore whether diluting samples with different ratios of water could improve the integrity of the signal in pooled results. While we tested samples with a range of different signal strengths, the detection of samples with even lower signals may warrant the use of smaller pools. Adding a few additional PCR cycles could be considered as a means to increase detection rate of such low viral load samples. In general, as RT-qPCR kits and protocols vary internationally, use of suggested pooling may require validation for each specific setting.

Due to technical limitations we pooled pure RNA for the RT-qPCR reaction, but it is also possible to pool samples prior to RNA extraction step. Doing so will also remove the emerging RNA extraction bottleneck. Additionally, while pooling at the RT-qPCR step does not allow running an internal control (endogenous gene), pooling prior to RNA extraction allows quality control for the RNA extraction step.

These results can be used not only for pooling, but also in multiplexing and any other signal compression techniques where samples are mixed to reduce the number of tests. We hope that this proof-of-concept will encourage others to develop mathematical and computational tools tailored for the pooling of SARS-CoV-2 tests.

Pooling is especially useful for routine community survey and for monitoring of cohesive groups. Local and global epidemic response critically depend on determining carriage frequency in the population, which is greatly enabled by pooling techniques. Furthermore, pooling techniques can be used for routine monitoring of essential work groups, such as hospital staff, military units, and factory workers. While the frequency of infection in these groups may be low, diagnosing even a single positive person typically requires quarantine of the entire group to prevent further spread in the community. In these surveillance applications, pooling may allow more routine monitoring and detection of low frequency of carriage thereby informing policy makers, reducing transmission, and alleviating the strain on healthcare services.

## Data Availability

All raw data is available in the supplementary figures.

---

**Supplementary Figure 1:**
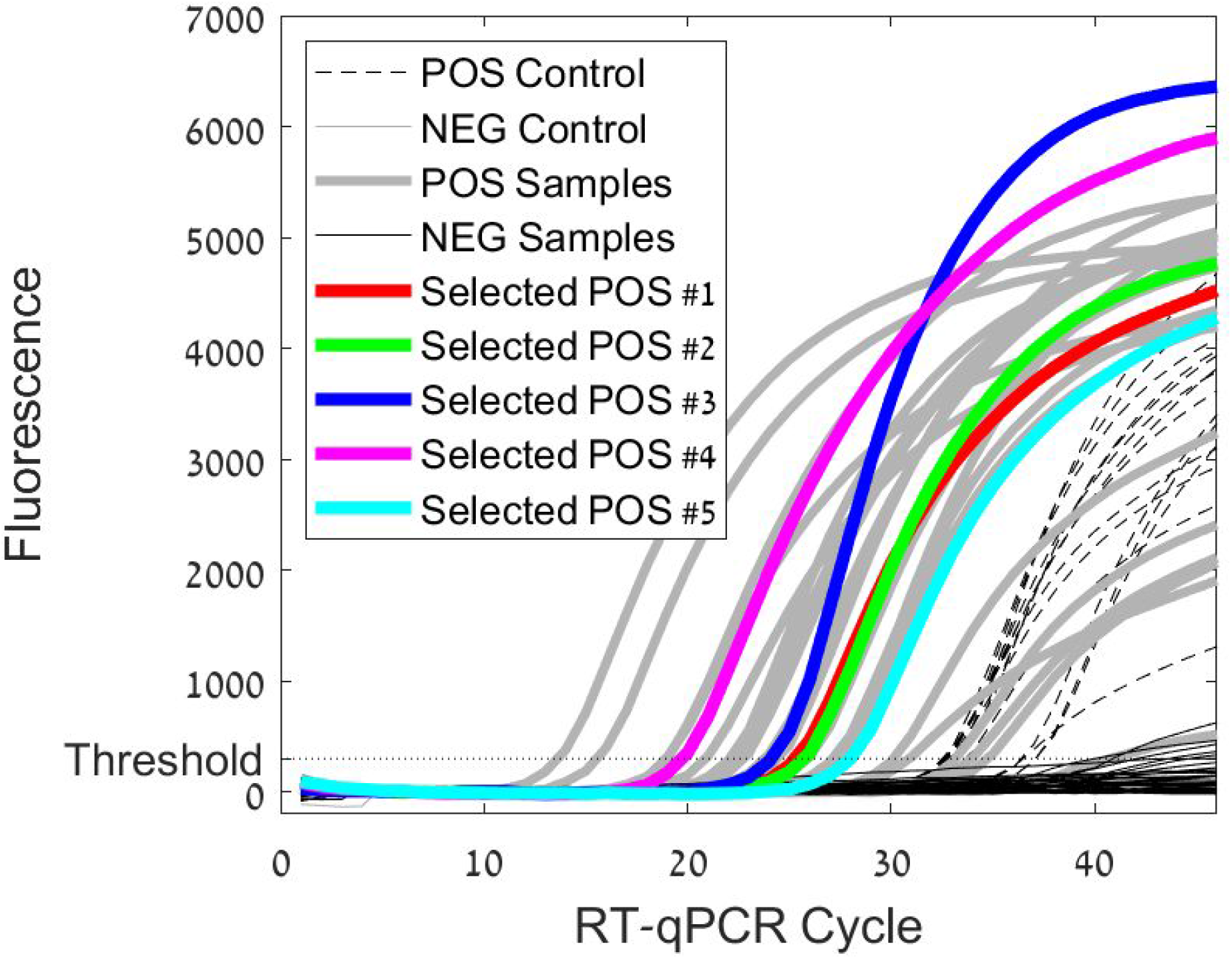
Selected Positive Samples are Representative of their Population Fluorescence of all samples as collected and tested at Rambam Health Care Campus between March 4-15. Selected positive samples are coloured (#1:red, #2:green, #3:blue, #4:pink, #5:turquoise) and are representative of all other positive-testing samples (grey). All positive samples and a few negative samples cross the threshold at a fluorescence of 300.

**Supplementary Figure 2:**
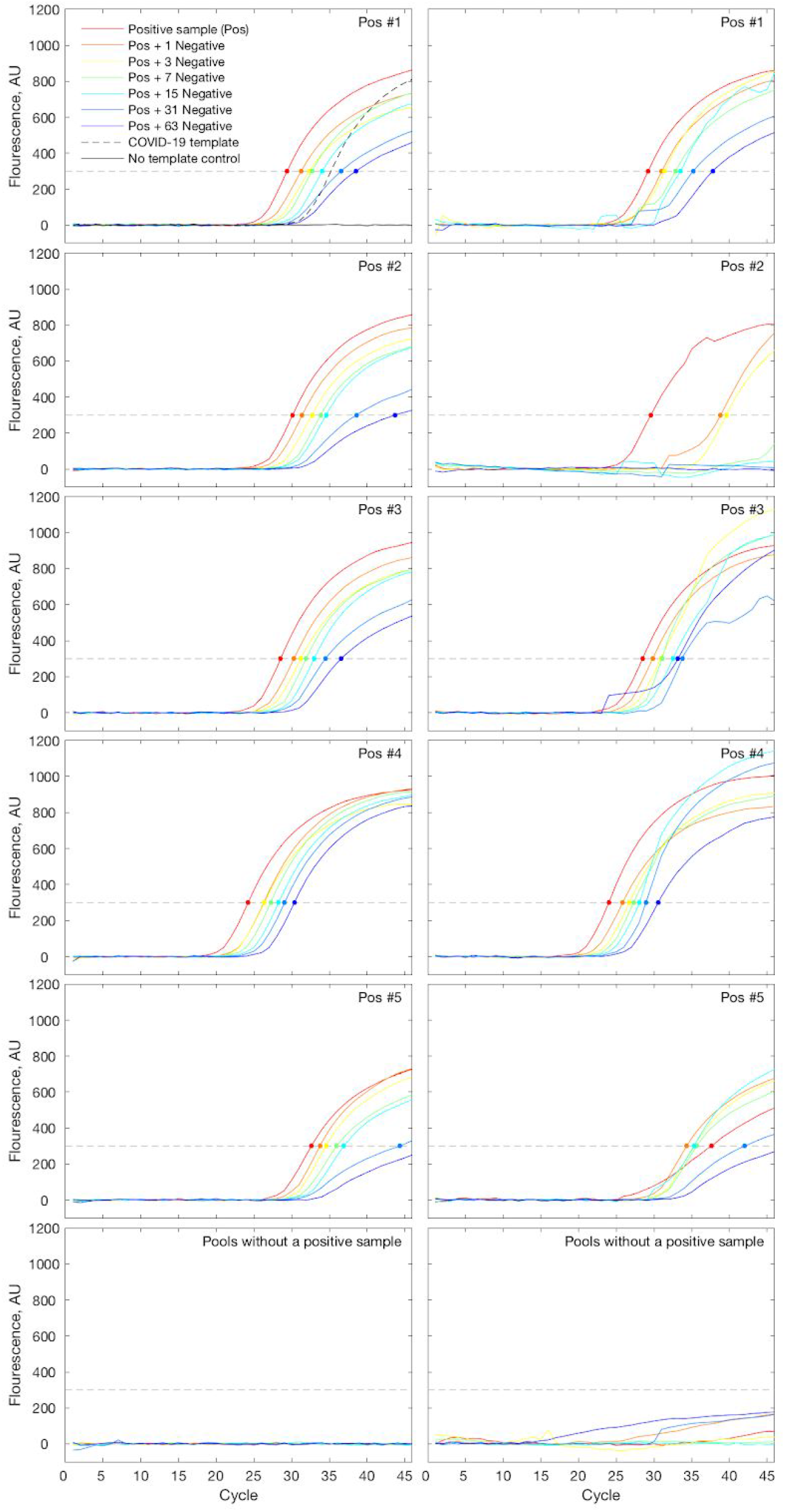
Fluorescence of pooled samples increases over Rt-qPCR cycles. Fluorescence over cycle during RT-qPCR for all positive samples (#1-#5 and control from up to down) over duplication A and B (left to right). Almost all pooled positive samples amplify in a sigmoidal curve that crosses the threshold.

